# Impaired cerebrovascular-cerebrospinal fluid coupling is associated with non-motor features of Lewy body disease

**DOI:** 10.64898/2026.05.13.26353090

**Authors:** Anna Ignatavicius, Ajay Konuri, Lachlan Churchill, Jack Anderson, Glenda M Halliday, Simon JG Lewis, Elie Matar

## Abstract

The temporal coupling between cortical blood-oxygen-level-dependent (BOLD) activity and CSF inflow has recently been proposed as a non-invasive marker of glymphatic function, a brain-wide clearance system closely linked to sleep, neuromodulatory regulation and neurodegeneration. Reduced BOLD-CSF coupling has been previously reported in Parkinson’s disease but its characterization in dementia with Lewy bodies, regional specificity and relevance to shared neuropsychiatric symptoms remain unclear. Using resting-state functional MRI, we quantified global and regional BOLD-CSF coupling in 39 participants, including 17 with Parkinson’s disease (mean age 61.4 years), 10 with dementia with Lewy bodies (mean age 72.8 years) and 12 healthy controls (mean age 66.2 years), and examined the relationship with clinical and cognitive measures, as well as volumetric measures of the subcortical ascending arousal network. Coupling was derived from the temporal cross-correlation of the global BOLD signal and CSF inflow timeseries. Group differences and associations with clinical, cognitive and volumetric measures of the subcortical ascending arousal network were assessed using non-parametric permutation testing. Parkinson’s disease and dementia with Lewy bodies patients both demonstrated weaker global BOLD-CSF coupling compared to controls, with no detectable difference between patient groups. Coupling reductions were most pronounced within the unimodal and attentional networks, encompassing regions that are particularly vulnerable in Lewy body disorders. Weaker coupling was associated with the severity of hallucinations and cognitive fluctuations, poorer nocturnal sleep quality and impaired attentional working memory, but not overall motor symptom burden. Associations between BOLD-CSF coupling and basal forebrain and brainstem volumes were observed, though partially age-dependent, suggesting a complex interaction between neuromodulatory system degeneration, ageing and brain-fluid dynamics. Our results provide preliminary evidence that disrupted temporal coordination between cerebrovascular activity and CSF inflow may contribute to the fluctuating neuropsychiatric features of Lewy body disorders and highlight the utility of BOLD-CSF coupling as a dynamic *in vivo* proxy of glymphatic function. Replication in larger cohorts incorporating multimodal imaging and biomarkers of pathology will be essential to validate these findings and determine whether brain-fluid dysregulation represents a potentially modifiable therapeutic target.

## Introduction

Lewy body disorders (LBD), including Parkinson’s disease (PD) and dementia with Lewy bodies (DLB), are α-synucleinopathies characterized by progressive motor, cognitive and neuropsychiatric symptoms. Although these disorders are typically distinguished by the temporal onset of parkinsonism relative to cognitive impairment, they share substantial clinical and pathological overlap, particularly with respect to non-motor symptoms.^1^ Visual hallucinations, cognitive fluctuations and sleep disturbances are disabling and ubiquitous in advanced PD and considered core diagnostic features of DLB.^2^ In the early stages of disease, these symptoms are often intermittent, suggesting the need for dynamic causal explanations, beyond structural neurodegeneration alone.

Considerable evidence points to alterations within the ascending arousal network (AAN) as a key contributing factor in their pathogenesis.^3-5^ The cholinergic and monoaminergic neurotransmitters associated with the subcortical AAN play an important role in regulating attention, arousal and sleep-wake transitions, exerting widespread effects on cortical activity and vascular tone.^6-8^ Recent experimental work has also implicated cholinergic and noradrenergic systems in coordinating neurovascular and cerebrospinal fluid (CSF) dynamics. Infraslow oscillatory noradrenergic locus coeruleus activity has been shown to modulate slow vasomotion during non-REM sleep, influencing CSF transport through the brain parenchyma,^9^ while ablation of basal forebrain cholinergic neurons disrupts the relationship between neurovascular coupling and CSF flux.^10^ Degeneration within the AAN may therefore contribute to symptoms of hallucinations, fluctuations and sleep disturbance via impaired cerebrovascular dynamics and CSF flow.

Sleep and arousal states have been tightly associated with the function of the glymphatic system,^11^ a highly organized brain network thought to facilitate the clearance of metabolic waste and pathogenic proteins through movement of CSF through perivascular spaces.^12^ While the exact mechanisms have yet to be completely understood, evidence from animal models has led to the prevailing theory that glymphatic transport is relatively enhanced during slow-wave sleep and may be mediated, in part, by low frequency spontaneous vasomotion and astrocytic regulation of perivascular flow.^11,13^ In humans, intrathecal contrast-enhanced MRI studies provide some support for a sleep-dependent relationship, showing that acute sleep deprivation impairs brain clearance.^14^ This process may therefore be affected by the sleep disturbance observed in PD and DLB.^5,15^ Consistent with this, diffusion tensor imaging along the perivascular space (DTI-ALPS), commonly interpreted as a structural biomarker of glymphatic system integrity,^16^ is reduced in LBD and has been associated with disease progression^17-19^ and phenoconversion from isolated REM sleep behavior disorder to PD.^20^ However, criticisms regarding the validity of DTI-ALPS as a direct measure of whole-brain glymphatic clearance,^21^together with subsequent reappraisals of its biological interpretation,^22^ suggest that it may be better understood as a spatially fixed-point marker capturing local tissue properties relevant to the brain-fluid environment. The complexity of the glymphatic system and broader brain clearance pathways highlights the need for complementary approaches that capture brain-wide and temporally dynamic aspects of neurofluid physiology.

Temporal coupling between global blood-oxygen-level-dependent (gBOLD) activity and CSF inflow has recently been proposed as a non-invasive surrogate marker of brain-fluid dynamics and glymphatic function.^23^ Under normal conditions, transient reductions in cerebral blood volume generate pressure gradients that drive compensatory CSF influx.^24^ First described in sleep,^25^ it has since been replicated during restful wakefulness in healthy individuals.^26^ Emerging evidence has revealed that gBOLD-CSF coupling alterations are present in both medicated and drug-naïve PD patients, and linked to disease progression,^27,28^ with similar findings reported in other neurodegenerative diseases.^23^ In parallel, there is also evidence to suggest that glymphatic clearance and brain-fluid coupling may be regionally specific. In Alzheimer’s disease (AD), a greater reduction of BOLD-CSF coupling strength has been observed in the anterior transmodal regions, consistent with early pathological changes involving the higher-order association networks.^29^ Regional specificity of BOLD-CSF coupling disturbances in LBD is unknown, however, complementary data suggest a unique pattern of posterior cerebrovascular vulnerability in LBD, including occipital and parietal hypometabolism and hypoperfusion,^30,31^ alongside altered functional connectivity of the visual and attentional networks.^32,33^ This raises the possibility that altered brain-fluid coupling may have a distinct spatial signature, with early and preferential involvement of unimodal sensory and attentional systems.

Hallucinations and cognitive fluctuations in LBD are closely linked to sleep disorders^34,35^ and their transient and state-dependent nature suggests that dynamic disturbances of cerebrovascular regulation and brain-fluid homeostasis may be especially relevant to their pathophysiology. At present, it remains unclear if disruption of BOLD-CSF coupling extends across the broader LBD spectrum, and whether such alterations relate to neuropsychiatric features associated with fluctuating vigilance and attentional dysfunction.

In this study, we examined global and regional BOLD-CSF coupling in PD and DLB and assessed the relationship between coupling strength and core clinical features of LBD. We hypothesized that BOLD-CSF coupling would be reduced in LBD compared to controls, and would be associated with cognitive status, hallucinations and sleep disturbance. To further understand the potential neuromodulatory basis of altered BOLD-CSF coupling, we explored associations with basal forebrain and brainstem AAN nuclei volume.

## Materials and methods

### Participants

Participants with PD and probable DLB were originally recruited and assessed at the Brain and Mind Centre, University of Sydney. Clinical diagnoses were made according to current consensus criteria.^2,36^ Healthy controls were recruited from the spouse population of the clinic and the local community. Control participants showed no evidence of cognitive impairment on formal neuropsychological testing and were screened to exclude major neurological or psychiatric disorders. All participants provided written informed consent in accordance with the Declaration of Helsinki, and the study was approved by the University of Sydney Human Research Ethics Committee (2013/HE000945).

### Clinical and neuropsychological assessment

Motor and non-motor symptoms were assessed using the Movement Disorder Society Unified Parkinson’s Disease Rating Scale (MDS-UPDRS).^37^ Sleep-related disturbances were assessed using the Scales for Outcomes in Parkinson’s Disease-Sleep (SCOPA-Sleep).^38^ The presence of hallucinations was defined based on a score ≥1 on MDS-UPDRS-I Q2. A subset of patients also completed the Psychosis and Hallucinations Questionnaire (PsycH-Q).^39,40^ The presence and severity of cognitive fluctuations in DLB were rated using the Clinician Assessment of Fluctuations Scale (CAF).^41^ Detailed cognitive assessment included the Montreal Cognitive Assessment (MoCA)^42^ to evaluate global cognition, together with measures of psychomotor speed and set-shifting (Trail Making Test Parts A and B), attention and working memory (Wechsler Memory Scale Digit Span Forward and Backward), and immediate and delayed episodic memory (Wechsler Memory Scale Logical Memory Parts I and II). Domain-specific cognitive scores were unavailable for four control participants and one DLB patient. Clinical assessment and MRI acquisition were performed while participants were on their usual medications. Additional details regarding clinical assessment are available in Supplementary Methods.

### MRI acquisition and preprocessing

All participants underwent a whole brain structural T1-weighted MRI scan and resting-state BOLD functional scan within six months of their clinical assessment. Imaging data were obtained on a 3-Tesla MRI scanner (General Electric). The total duration of the resting-state scan was approximately 7 minutes during which participants were instructed to lie awake with their eyes closed.

Preprocessing of fMRI data was performed using fMRIPrep 24.1.1.^43^ Major preprocessing steps included motion and intensity non-uniformity correction, co-registration, normalization, resampling for spatial alignment across participants and confound estimation. T1-weighted images were preprocessed using FreeSurfer version 7.3.2 with the standard "recon-all" pipeline for whole-brain segmentation. Visual quality control reports were generated for each participant to ensure preprocessing accuracy. A detailed description of acquisition parameters and preprocessing pipeline is provided in the Supplementary Methods.

### Global BOLD and CSF inflow signal extraction

The gBOLD signal was derived by averaging voxelwise BOLD timeseries across gray matter regions defined using the Harvard-Oxford structural atlas,^44^ warped from MNI152NLin6Asym standard space to each participant’s native BOLD space (Supplementary Fig S1). CSF inflow timeseries were extracted from a manually delineated mask placed at the most inferior slice of the raw functional image, where inflow-related intensity changes are expected to be maximal.^25^ CSF voxels were identified based on increased signal intensity compared to surrounding tissue on T2*-weighted functional images and confirmed with co-registered T1-weighted structural scans (Supplementary Fig S1). Only participants with a clearly identifiable CSF inflow signal within the functional field of view were included for formal analysis. Further details are provided in Supplementary Methods.

### Quantification of BOLD signal and CSF inflow coupling

Global BOLD-CSF coupling was computed using cross-correlation functions between the gBOLD signal and the CSF signal across time lags of ±18 s (±6 TRs).^25,27^ Averaged group-level peak negative lag was identified and used to quantify the strength of coupling for each participant. Coupling was additionally assessed by calculating the cross-correlation functions between the first-order negative temporal derivative of the gBOLD signal and the CSF signal.^25,26^ Statistical significance was assessed using permutation resampling (10000 iterations), which involved randomly reassigning gBOLD and CSF signals across participants to generate a null distribution.

To examine spatial variation in BOLD-CSF coupling across the cortex, the Schaefer-400 functional atlas^45^ was transformed from MNI space into each participant’s native functional space and voxelwise BOLD time series were averaged within each cortical parcel. For each parcel, the BOLD-CSF coupling was quantified as the cross-correlation value at the parcel-specific negative peak.^46^ Parcelwise coupling estimates were first averaged within canonical resting-state functional networks according to the Yeo 7-network scheme.^47^ Network-level coupling was subsequently summarized into unimodal (visual and somatomotor networks), attentional (dorsal and ventral attention networks), and transmodal (frontoparietal and default mode networks) systems, following the principal gradient of macroscale cortical functional organization.^48^ Supplementary analyses additionally assessed group differences in coupling within each individual functional network.

### Basal forebrain and brainstem volume estimation

Basal forebrain (BF) and brainstem nuclei were derived from T1-weighted images using automated segmentation toolboxes designed for integrated use with FreeSurfer. The BF was segmented using ScLimbic,^49^ a deep-learning-based tool for the automatic segmentation of subcortical limbic structures. Brainstem nuclei volumes corresponding with the AAN were extracted using AANSegment,^50^ a recently developed automated Bayesian segmentation algorithm that uses a probabilistic atlas derived from *ex vivo* brain specimens with corresponding histopathological annotations. Segmentations were performed in native T1-weighted structural MRI space and visually inspected for quality control. Basal forebrain and whole brainstem AAN volumes were exported from segmentation outputs in mm³. Total gray matter volume and bilateral striatal volume were additionally extracted for supplementary analyses examining broader and disease-relevant structural associations. All volumes were adjusted for total intracranial volume using residual correction. Further segmentation details are provided in the Supplementary Methods.

### Statistical analysis

Analyses were performed in MATLAB R2023a. Group differences were assessed using non-parametric permutation testing (10000 permutations). Planned pairwise contrasts comparing global and regional BOLD-CSF coupling between diagnostic groups included age and sex as covariates, with inference based on the two-tailed permutation distribution of the signed t-statistic. Associations with clinical, cognitive and volumetric measures were examined using permutation-based regression models, controlling for age and sex, with additional adjustment for education for analyses of cognitive performance. Neuropsychological test scores were converted to z-scores prior to analysis.

Effect sizes are reported as Hedges’ *g* for group comparisons and standardized regression coefficients (β) for association analyses. Statistical significance was set at *p* < 0.05, with 95% confidence intervals (CI) estimated using bootstrapping (5000 resamples). Multiple comparisons for pre-specified analyses were controlled using false discovery rate (FDR) correction. Exploratory analyses were conducted without correction where explicitly stated. Robustness of exploratory regression findings was assessed using jackknife leave-one-out (LOO) sensitivity analyses, in which regression models were iteratively re-estimated after excluding one participant at a time.

## Results

### Demographics

This study included a total of 39 participants (12 controls, 17 PD and 10 DLB). Compared with both PD patients and controls, DLB patients were significantly older and demonstrated poorer global cognitive performance. DLB patients also exhibited greater overall motor and non-motor symptom burden compared to PD patients. At the time of assessment, seven PD (41%) and eight DLB patients (80%) reported hallucinations. Cognitive fluctuations were reported in seven DLB patients (70%). As expected, all PD patients were receiving dopaminergic therapy, and the majority of DLB patients (70%) were receiving cholinesterase inhibitor treatment. Details regarding demographic and clinical characteristics are outlined in Table 1.

**Table 1.** Participant characteristics.

|  | **Controls (n = 12)** | **PD (n = 17)** | **DLB (n = 10)** | **p** |
| --- | --- | --- | --- | --- |
| Age (years) | 66.2 ± 7.13 | 61.4 ± 9.65 | 72.8 ± 5.18<sup>b,c</sup> | 0.004 |
| Sex (male, n %) | 2 (17%) | 10 (58%) | 8 (80%) | - |
| Education (years) | 14.3 ± 2.67 | 13.9 ± 2.85 | 12.5 ± 3.44 | 0.358 |
| MoCA | 27.8 ± 1.47 | 27.5 ± 2.45 | 19.5 ± 5.20<sup>b,c</sup> | <0.001 |
| Disease duration (years) | - | 5.36 ± 3.86 | 1.25 ± 2.27 | 0.007 |
| MDS-UPDRS I | - | 8.53 ± 3.57 | 18.1 ± 7.06 | 0.002 |
| MDS-UPDRS II | - | 9.18 ± 6.56 | 16.2 ± 6.49 | 0.012 |
| MDS-UPDRS III | - | 21.2 ± 12.8 | 32.4 ± 10.8 | 0.028 |
| SCOPA-Sleep Night | - | 5.65 ± 3.81 | 3.00 ± 3.94 | 0.096 |
| SCOPA-Sleep Day | - | 4.47 ± 2.62 | 7.40 ± 3.41 | 0.020 |
| Hallucinations (n %) | - | 7 (41%) | 8 (80%) | - |
| MDS-UPDRS Hallucinations | - | 0.65 ± 1.00 | 1.70 ± 1.42 | 0.036 |
| Fluctuations (n %) | - | - | 7 (70%) | - |
| ChEI (% on) | - | 0 (0%) | 7 (70%) | - |
| Levodopa (% on) | - | 17 (100%) | 4 (40%) | - |
| LEDD (mg/day) | - | 671 ± 450 | 350 ± 191 | 0.188 |

Data are expressed as *n* (%) or mean and standard deviation. Disease duration was calculated as the time interval between clinical diagnosis and MRI acquisition. Levodopa daily dose equivalents were calculated according to standard conversion formulas (mg/day). P values were calculated using non-parametric permutation testing (10000 permutations). <sup>a</sup>Pairwise comparisons: significant differences between PD and controls; <sup>b</sup>Pairwise comparisons: significant differences between DLB and controls; <sup>c</sup>Pairwise comparisons: significant differences between PD and DLB.
ChEI, cholinesterase inhibitor; DLB, dementia with Lewy bodies; LEDD, levodopa equivalent daily dose; MDS-UPDRS, Movement Disorder Society Unified Parkinson's Disease Rating Scale; MoCA, Montreal Cognitive Assessment; PD, Parkinson's disease; SCOPA-Sleep, Scale for Outcomes in Parkinson's Disease-Sleep.

### Global BOLD-CSF coupling is altered in LBD

To determine whether the gBOLD activity was temporally coupled to CSF inflow, cross-correlation functions between the gBOLD signal and CSF signal were computed and averaged across all participants. Consistent with previous reports,^25,27^ the mean gBOLD-CSF cross-correlation function demonstrated a positive peak at a lag of -6 seconds (*r* = 0.250, *p* < 0.001) and a negative peak at a lag of +3 seconds (*r* = −0.244, *p* < 0.001; Fig. 1A). The cross-correlation function between the CSF signal and the negative first-order derivative of the gBOLD signal showed a large positive peak around a lag of -3 seconds (*r* = 0.243, *p* < 0.001; Fig. 1B). Therefore, the gBOLD-CSF coupling strength for each participant was quantified as the cross-correlation value at +3 seconds.

**Figure 1.**
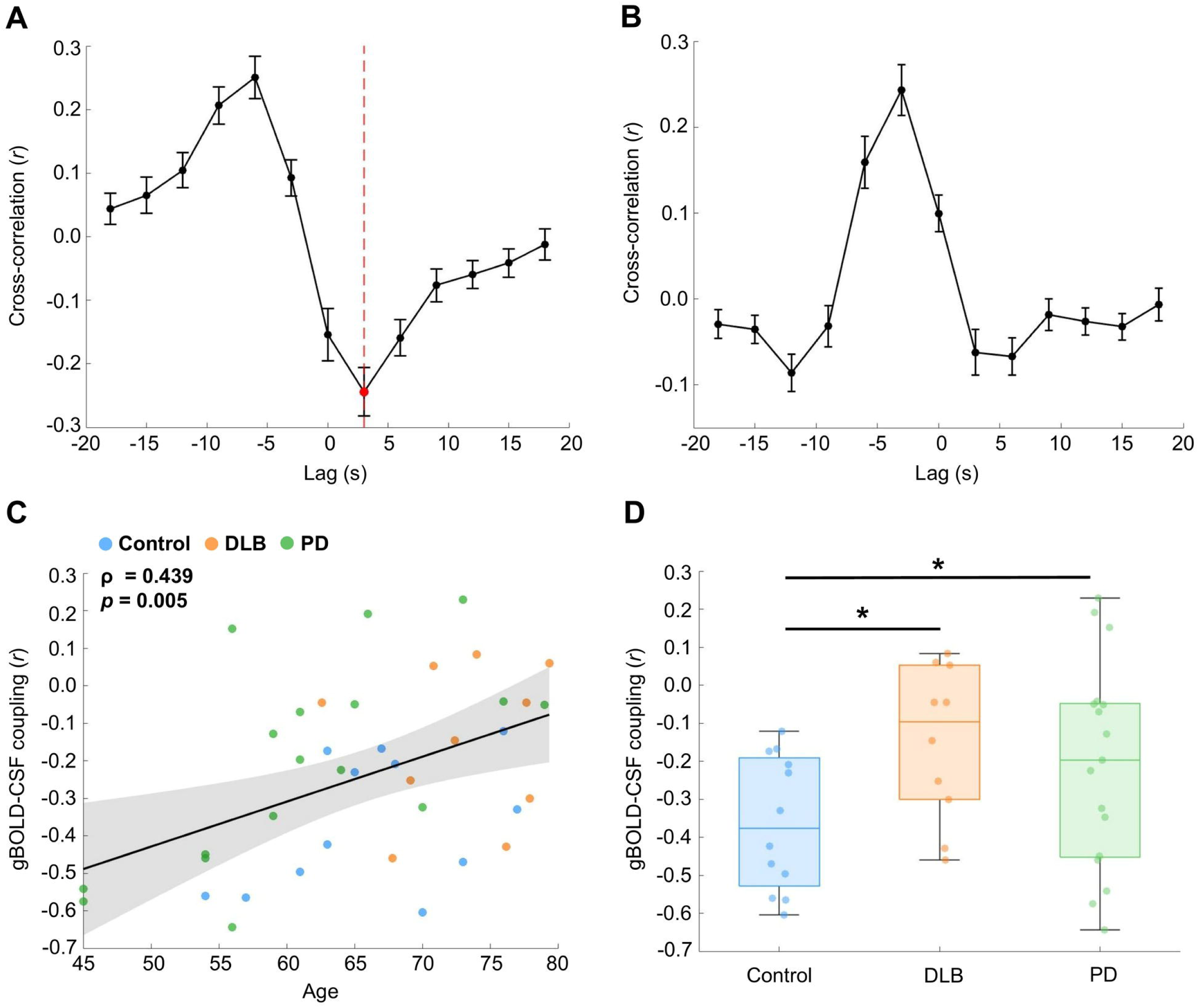
Global BOLD-CSF coupling is associated with age and is reduced in Lewy body disease. The group averaged cross-correlation function between global BOLD signal and CSF signal across the whole sample (*n* = 39), showed a positive peak at the -6s lag (mean *r* = 0.250, *p* < 0.001, 10000 permutations) and a negative peak (red dashed line) at the +3s lag (mean *r* = -0.244, *p* < 0.001, 10000 permutations; **A**). The cross-correlation between CSF signal and the negative first-order derivative of the global BOLD signal showed a positive peak at the -3s lag (*r* = 0.243, *p* < 0.001, 10000 permutations; **B**). The group-derived +3s lag was used to quantify individual global BOLD-CSF coupling strength. Global BOLD-CSF coupling strength decreased (shift away from strong anti-correlation) with increasing age (**C**). Both Parkinson’s disease (PD) and dementia with Lewy bodies (DLB) patients demonstrated weaker gBOLD-CSF coupling compared to controls, controlling for age and sex (**D**). Error bars represent the standard error of the mean. Box plots show median and interquartile range; data points represent individual participants. Asterisks demonstrate FDR-corrected *p* < 0.05.

At the individual level, increasing age was associated with weaker gBOLD-CSF coupling (ρ = 0.439, *p* = 0.005; Fig 1C). Compared to controls, both PD and DLB patients demonstrated reduced gBOLD-CSF coupling strength (PD: Hedges’ *g* = 0.774, *p_FDR_* = 0.037; DLB: Hedges’ *g* = 0.660, *p_FDR_* = 0.037; Fig 1D). No differences were observed between PD and DLB (*p_FDR_* = 0.556).

### Reduced global BOLD-CSF coupling is linked to sleep disturbance, hallucinations and cognitive fluctuations

Within the combined LBD patient group (pooled PD and DLB participants), weaker gBOLD-CSF coupling strength was associated with higher SCOPA-Sleep Nocturnal scores (β = 0.631, bootstrap 95% CI [0.257, 0.843], *p_FDR_*= 0.005; Fig. 2A), as well as higher scores on the hallucinations item of the MDS-UPDRS (β = 0.533, bootstrap 95% CI [0.306, 0.754], *p_FDR_* = 0.015; Fig. 2B), after adjusting for age and sex. No associations were observed between gBOLD-CSF coupling and MDS-UPDRS scores or SCOPA-Sleep Day scores (*p* > 0.05, Supplementary Table S1).

**Figure 2.**
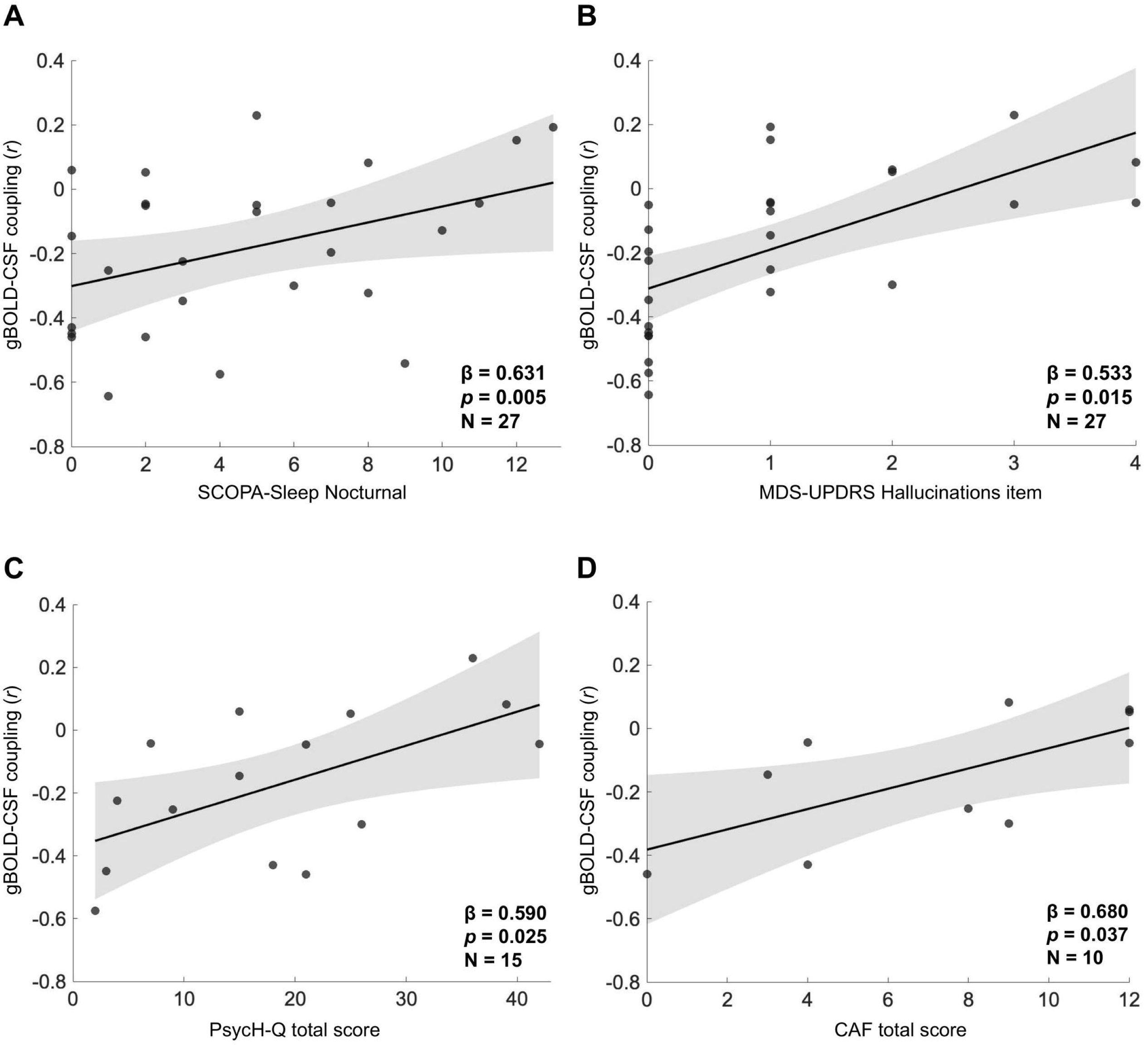
Global BOLD-CSF coupling strength is associated with core clinical features of Lewy body disease. Weaker global BOLD-CSF coupling was associated with poorer subjective nocturnal sleep (SCOPA-Sleep Night; **A**) and hallucination severity (MDS-UPDRS Hallucinations item; **B**) after controlling for age and sex. Subset analyses demonstrated associations between weaker global BOLD-CSF coupling and increased burden of psychosis-related symptoms (PsycH-Q; **C**) and cognitive fluctuation severity (CAF; **D**). Scatter plots display raw data; each data point represents an individual participant. Shaded region represents 95% confidence interval of the regression line. Statistics derived from permutation-based regression models (two-sided, 10000 permutations); FDR corrected for panels A and B. PsycH-Q and CAF results (C, D) are exploratory and uncorrected for multiple comparisons.

In exploratory subset analyses, weaker gBOLD-CSF coupling strength was also associated with higher scores on the PsycH-Q (*n* = 15 [DLB: 10, PD: 5]; β = 0.590, bootstrap 95% CI [0.179, 0.832], *p* = 0.025; Fig. 2C) and greater cognitive fluctuation severity in DLB patients (*n* = 10; β = 0.680, bootstrap 95% CI [0.191, 0.917], *p* = 0.037, Fig. 2D). Sensitivity analyses using jackknife LOO resampling demonstrated that the direction of association remained consistent across all iterations for both PsycH-Q and CAF, suggesting that the observed associations were not driven by single influential observations (Supplementary Table S2 and Supplementary Fig. S2).

### Global BOLD-CSF coupling strength is associated with domain-specific cognition

After controlling for age and sex, no association between gBOLD-CSF coupling and global cognition as measured by MoCA was observed, either across the whole sample or within the LBD patient cohort (both *p* > 0.05, Supplementary Table S3). However, reduced gBOLD-CSF coupling was associated with poorer attentional working memory reflected by lower Digit Span Backwards scores (β = -0.577, bootstrap 95% CI [-0.754, -0.367], *p_FDR_* = 0.004; Fig. 3A). This association remained significant when restricted to LBD patients (β = -0.550, bootstrap 95% CI [-0.782, -0.251], *p_FDR_* = 0.035).

**Figure 3.**
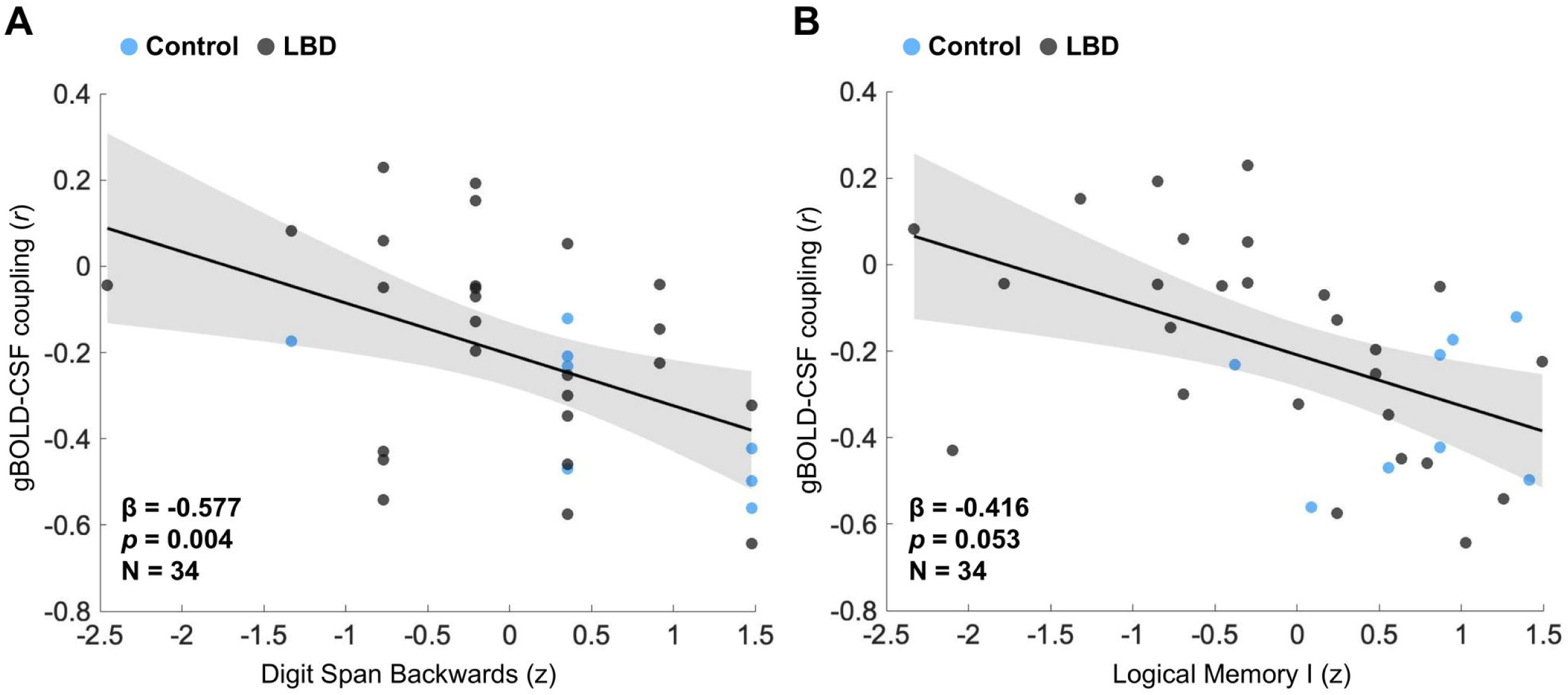
Associations between global BOLD-CSF coupling strength and domain-specific cognitive performance. Weaker global BOLD-CSF coupling strength was associated with poorer working memory performance (Digit Span Backwards; **A**) with a trend-level association between weaker gBOLD-CSF coupling and immediate episodic memory performance (Logical Memory Part I; **B**). Scatter plots display raw data; each data point represents an individual participant. Blue points denote control participants and black points denote the combined Lewy body disorder (LBD) cohort (pooled Parkinson’s disease and dementia with Lewy bodies participants). Shaded region represents 95% confidence interval of the regression line. Statistics derived from permutation-based regression models (two-sided, 10000 permutations) adjusting for age, sex and education (FDR corrected).

Within the whole sample, there was also evidence for a relationship between weaker gBOLD-CSF coupling and worse immediate episodic memory (Logical Memory Part I: β = -0.416, bootstrap 95% CI [-0.730, -0.090], *p* = 0.015; Fig. 3B), although this did not survive correction for multiple comparisons (*p_FDR_* = 0.053).

### Selective reduction of BOLD-CSF coupling across the functional hierarchy in LBD

To test whether disease-related alterations in BOLD-CSF coupling preferentially affected specific levels of the cortical functional hierarchy, coupling strength was examined across unimodal, attentional and transmodal systems. Compared to controls, combined LBD patients exhibited weaker BOLD-CSF coupling in both the unimodal brain networks (Hedges’ *g* = 0.831, *p_FDR_* = 0.021), and the attentional brain networks (Hedges’ *g* = 0.648, *p_FDR_* = 0.048), with only a marginal difference observed in the transmodal networks (Hedges’ *g* = 0.501, *p* = 0.077). No group differences in regional BOLD-CSF coupling were observed between PD and DLB (*p* > 0.05). Individual network comparisons demonstrated significant group differences in visual, somatomotor and dorsal attention networks, consistent with the primary finding of preferential involvement of unimodal and attentional systems (Supplementary Table S4). Figure 4 illustrates the cortical expression of BOLD-CSF coupling for each group, and the spatial pattern of reduced coupling in LBD relative to controls.

**Figure 4.**
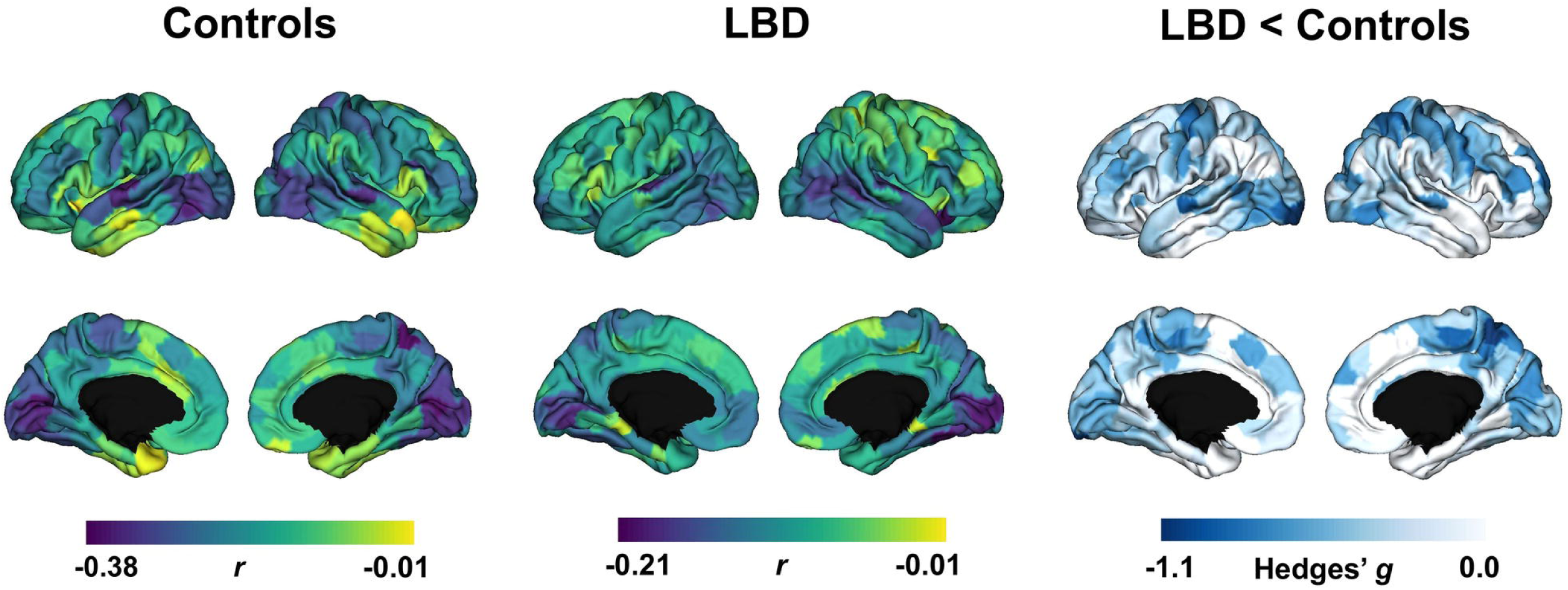
Cortical expression of BOLD-CSF coupling and regional reductions in Lewy body disease. Surface maps illustrating parcel-wise BOLD-CSF coupling strength across the cortex in healthy controls (left panel; n = 12), combined Lewy body disorder (LBD) patients (middle panel; n = 27; Parkinson’s disease n = 17, dementia with Lewy bodies n = 10), and the spatial distribution of relative coupling reductions in LBD compared with controls, highlighting a pattern that predominantly involves the unimodal and attentional networks (right panel). Color bars indicate parcel-wise coupling strength (*r*) for each group and effect size (Hedges’ *g*) for between group comparisons. Parcel-wise Hedges’ g values are shown for visualization of spatial topography; formal statistical inference on regional coupling was restricted to the three hierarchical network composites (unimodal, attentional, transmodal; see Results). Data are displayed on the lateral and medial cortical surface.

### Association of BOLD-CSF coupling with basal forebrain and brainstem ascending arousal network volumes

In the entire sample, larger BF volumes and brainstem AAN volumes were associated with stronger (more negative) gBOLD-CSF coupling (BF: β = -0.384, bootstrap 95% CI [-0.626, - 0.101], *p_FDR_* = 0.019; brainstem AAN: β = -0.431, bootstrap 95% CI [-0.636, -0.182], *p_FDR_* = 0.009). Similar directionality and magnitude of effects were observed within the combined LBD group (BF: β = −0.458, bootstrap 95% CI [-0.725, -0.106], *p_FDR_* = 0.018; brainstem AAN: β = −0.451, bootstrap 95% CI [-0.672, -0.146], *p_FDR_*= 0.018). Following adjustment for age and sex, associations attenuated, with the relationship between brainstem AAN volume and gBOLD-CSF coupling in the whole sample remaining at trend level (β = -0.316, bootstrap 95% CI [-0.546, -0.037], *p* = 0.052, *p_FDR_* = 0.104).

We next examined associations between regional BOLD-CSF coupling within the unimodal and attentional systems, as these networks demonstrated altered coupling within LBD. In the whole sample, larger brainstem AAN volume was associated with stronger BOLD-CSF coupling within unimodal (β = -0.348, bootstrap 95% CI [-0.559, -0.093], *p* = 0.031) and attentional networks (β = -0.358, bootstrap 95% CI [-0.561, -0.124], *p* = 0.027) after adjustment for age and sex, however, neither survived correction for multiple comparisons (*p_FDR_* = 0.093).

Within LBD patients, larger BF volume was associated with stronger unimodal and attentional BOLD-CSF coupling (β range: -0.462 to -0.521; *p_FDR_* range: 0.015-0.018). Similarly, larger brainstem AAN volume was also associated with stronger unimodal and attentional BOLD-CSF coupling (β range: -0.485 to -0.523; *p_FDR_* range: 0.015-0.018). However, these associations were no longer significant after covarying for age and sex. A full summary of results is provided in Table S5 in Supplementary Results.

Total gray matter volume was associated with stronger BOLD-CSF coupling across global, unimodal and attentional measures in unadjusted models, both in the whole sample and within the LBD subgroup (Supplementary Table S6). These associations were attenuated after adjustment for age and sex and did not survive FDR correction. In contrast, striatal volume was not significantly associated with global or regional BOLD-CSF coupling in either the whole sample or the LBD subgroup, in unadjusted or age- and sex-adjusted models (Supplementary Table S6).

### Additional control analyses

Head motion, indexed by mean framewise displacement (FD) and the proportion of high-motion volumes, did not differ significantly between diagnostic groups (all p > 0.05; Supplementary Table S7) and was not associated with either global or regional BOLD-CSF coupling (all p > 0.05). Group comparisons between controls and LBD patients including mean FD as an additional covariate showed consistent direction and magnitude of effect for both global and regional BOLD-CSF coupling (Supplementary Table S8). Associations between levodopa equivalent daily dose (LEDD) and BOLD-CSF coupling were also examined among levodopa-treated participants. LEDD was not associated with global or regional BOLD-CSF coupling in either PD alone or the combined LBD subgroup receiving dopaminergic therapy (all *p* > 0.05, Supplementary Table S9).

## Discussion

In this study, we demonstrated altered temporal coupling of global hemodynamic activity and CSF inflow in both PD and DLB relative to healthy controls, with preferential disruption within unimodal and attentional systems. Importantly, weaker gBOLD-CSF coupling was associated with hallucinations, cognitive fluctuations, poorer subjective sleep quality and working memory impairment but not motor symptom severity, suggesting relative specificity to the cognitive and neuropsychiatric manifestations of LBD. Structural alterations to the basal forebrain and brainstem AAN were also related to reduced BOLD-CSF coupling, although these associations were partially age-dependent. Collectively, these results suggest that the region-specific decoupling of cerebrovascular and CSF dynamics may represent a systems-level substrate linked to arousal instability and paroxysmal symptoms of LBD.

Our findings build upon prior reports of weaker BOLD-CSF coupling in PD,^27,28^ and uniquely extend these observations to DLB, which has thus far been minimally characterized using dynamic measures of brain-fluid physiology. The absence of detectable differences between patient groups suggests that impaired cerebrovascular-CSF coordination may be a shared pathophysiological feature across the LBD spectrum. We also observed that weaker BOLD-CSF coupling in LBD was not spatially uniform but was most pronounced within the unimodal and attentional networks, encompassing regions that are particularly vulnerable in PD and DLB. Posterior hypoperfusion is a distinguishing feature of LBD^51^ and is detectable across a range of modalities.^30,31,52,53^ *In vivo* evidence suggests that this could reflect local neurovascular dysregulation, with recent studies finding impaired coupling between generalized cerebral blood flow and regional functional connectivity as well as delayed cerebrovascular reactivity within the sensory association cortices.^54,55^ As such, impaired neurovascular responses could disrupt the coordination between transient cerebral blood flow and CSF influx.^15^ Supporting this, a recent post-mortem study in PD identified alterations to both the microvasculature and key cellular components of the glymphatic system within the occipital cortex, despite limited Lewy body pathology.^56^ These changes included collapsed capillary beds, pericyte loss, enlarged perivascular spaces and reduced localization of aquaporin-4 (AQP4) water channels to astrocytic endfeet, which are essential structures regulating solute exchange between the CSF and interstitial fluid.^12,13^ Together, these findings suggest that the more severe reduction in BOLD-CSF coupling within sensory and attentional networks may be a physiological signature linking regional neurovascular dysfunction with compromised brain-fluid homeostasis in LBD.

The spatial pattern in LBD contrasts with that reported in AD, where disruptions of BOLD-CSF coupling have been shown to preferentially affect higher-order transmodal networks, corresponding with the regional distribution of early amyloid deposition and functional connectivity changes.^29^ Reduced connectivity within the default mode network (DMN) is a characteristic feature of AD, apparent even in asymptomatic and prodromal stages.^57,58^ In PD and DLB, however, findings regarding functional alterations within the DMN have been more heterogeneous,^32,59-61^ likely reflecting variable degrees of concomitant Alzheimer-type pathology. Indeed, greater cortical amyloid and tau burden in DLB has been associated with a more pronounced loss of posterior DMN connectivity.^58^ These observations suggest that brain-fluid disruption in LBD may initially affect the sensory and attentional systems, with transmodal network involvement coinciding with the presence of mixed pathology. Longitudinal studies incorporating imaging markers of amyloid and tau will be important for clarifying how co-pathology relates to the spatial and temporal progression of altered BOLD-CSF coupling in LBD.

Converging evidence from neuropathological, neuroimaging and preclinical studies suggests that impaired glymphatic function may contribute to neurodegenerative processes underlying LBD.^15,56,62,63^ Although not a direct measure of glymphatic transport, experimental work has demonstrated that BOLD-CSF coupling correlates with tracer-based measures of tissue glymphatic signal kinetics.^10^ Consistent with the established relationship between sleep and glymphatic clearance, we found that weaker gBOLD-CSF coupling was associated with poorer subjective nighttime sleep quality in LBD, suggesting disrupted nocturnal sleep may impair brain-fluid dynamics in a manner that persists into wakefulness. This finding aligns with a previous study in PD that demonstrated reduced gBOLD-CSF coupling in patients with more severe sleep-related symptoms.^28^ Prior work has shown a dissociation of gBOLD-CSF coupling responses to sleep deprivation between young adults (increased coupling) and older adults (reduced coupling),^64^ consistent with the well-documented age-related decline in glymphatic function^11^ and broader evidence linking sleep disturbance to neurodegenerative risk.^65^ Age is an important modifier of disease course in LBD, and a predictor of cognitive impairment and more rapid disease progression.^1,66^ In our findings, the association between poorer sleep quality and reduced coupling persisted after adjusting for age, indicating additive disease-specific dysfunction in LBD.

Recent work has further demonstrated that attentional lapses following sleep deprivation occur in temporal proximity to global neurovascular events and pulsatile waves of CSF flow, suggesting a relationship between brain-fluid dynamics and moment-to-moment cognitive stability.^67^ In this context, our finding that weaker gBOLD-CSF coupling was associated with greater severity of cognitive fluctuations and hallucinations suggests that disrupted cerebrovascular-CSF coordination may contribute to paroxysmal symptoms of LBD related to altered arousal and attentional processing.^3,4^ Previous studies have highlighted a role for aberrant whole brain dynamics underlying these phenomena, with dysfunctional regions mapped to areas with high expression of cholinergic and noradrenergic receptor genes.^68,69^

Cholinergic and noradrenergic systems operate across diverse spatiotemporal scales and are thought to modulate dynamic network topology and brain state transitions by altering neural gain.^70,71^ Emerging evidence also implicates both the cholinergic nucleus basalis of Meynert and the noradrenergic locus coeruleus in the regulation of neurovascular and glymphatic function.^9,10^ Altered BOLD-CSF coupling and the cognitive and psychotic features of LBD may therefore reflect a shared downstream consequence of dysfunction within these arousal systems. More speculatively, impaired CSF and interstitial fluid exchange may affect volume transmission of acetylcholine and noradrenaline acting through extracellular and perivascular spaces.^72,73^ In this manner, decoupling of CSF flow from global hemodynamic oscillations could also compromise the balance of neuromodulatory tone relevant to maintaining coordinated shifts between brain states, exacerbating existing vulnerabilities in these systems.

Glymphatic dysfunction likely reflects a complex interplay of age-related and neurodegenerative alterations to sleep-wake circuitry, cerebrovascular regulation and neuromodulatory systems. Although basal forebrain and brainstem AAN volumes were associated with BOLD-CSF coupling, these relationships attenuated after adjustment for age and sex. Both subcortical AAN volumes and BOLD-CSF coupling decline with advancing age, therefore it is difficult to disambiguate disease-specific effects from age-related processes, especially in LBD where older age may additionally reflect greater accumulated co-pathology.^1^ Furthermore, while aggregation of α-synuclein occurs within these regions early in PD and DLB, the functional consequences of these inclusions may precede overt neuronal loss,^71,74,75^ suggesting that volumetric measures alone may underestimate early neuromodulatory disruption.

### Limitations and future research

There are several limitations that should be considered. Quantification of BOLD-CSF coupling required adequate inferior field of view coverage to reliably capture CSF inflow at the lower boundary of the acquisition volume, which constrained the available sample size. Although significant associations between reduced BOLD-CSF coupling and important clinical features of LBD were observed, our sample may have been underpowered to detect smaller yet biologically meaningful effects therefore these findings warrant replication and validation in larger cohorts. Differences in medication exposure between groups, including potential effects of cholinesterase inhibitor use,^10^ may have contributed to variability in coupling strength and potentially influenced the absence of measurable differences between PD and DLB. Carefully controlled studies in drug-naïve or medication-stratified cohorts will be essential to disentangle intrinsic disease effects from potential pharmacological modulation of neurovascular-CSF dynamics. Both oscillations of the BOLD signal and CSF inflow are sensitive to physiological responses unrelated to neural activity, including cardiorespiratory pulsations.^23^ Our scan time was kept relatively short, however, we cannot exclude the influence of inter-individual differences in arousal level and autonomic factors on BOLD-CSF coupling strength. Incorporating multimodal physiological monitoring may help to parse out the distinct neural and autonomic contributions to BOLD-CSF coupling in LBD. Due to their small and complex anatomical boundaries, we evaluated the basal forebrain and brainstem nuclei as composite regions, which may have obscured subtle nucleus-specific alterations and their unique contribution to BOLD-CSF coupling. Integration of high-resolution MRI, neuromelanin-sensitive sequences, and neurotransmitter-specific molecular imaging will be needed to further clarify the role of these structures. Lastly, the cross-sectional nature of this study means we cannot determine the temporal relationship between altered BOLD-CSF coupling and α-synuclein pathology. Longitudinal studies in prodromal and early disease cohorts, together with further development of α-synuclein-specific *in vivo* biomarkers, may help to elucidate how disrupted cerebrovascular-CSF dynamics relate to synucleinopathy progression.

## Conclusion

In summary, our findings demonstrate that the temporal coupling between cerebrovascular activity and CSF inflow is altered in LBD and provide preliminary evidence of a potential mechanism linking sleep disturbance, brain-fluid dysregulation and paroxysmal non-motor features of LBD. Future investigations in larger cohorts integrating BOLD-CSF coupling with electrophysiological, structural and molecular biomarkers will be essential to establish whether disrupted brain-fluid dynamics represents a tractable therapeutic target in LBD.

## Data availability

The derived source data and code used in this study are listed in a Key Resource Table alongside their persistent identifiers at https://doi.org/10.5281/zenodo.21897287.

Detailed protocols for clinical and neuropsychological assessment (https://doi.org/10.17504/protocols.io.8epv5w336v1b/v1) and MRI acquisition and preprocessing (https://doi.org/10.17504/protocols.io.8epv5w3b6v1b/v1) are publicly available through protocols.io.

Deidentified individual-level raw data are available from the corresponding author upon reasonable request. Access is subject to confirmation of appropriate institutional ethics approval and completion of a Material Transfer Agreement before data transfer.

An earlier version of this manuscript was posted to MedRxiv on 2026-08-17 at https://doi.org/10.64898/2026.05.13.26353090

