## Supplementary Material, Table S1, Fig. S1 for "Impaired cerebrovascular-cerebrospinal fluid coupling is associated with non-motor features of Lewy body disease"

### 1. Supplementary methods

#### 1.1 Clinical and neuropsychological assessment

Comprehensive medical history was obtained via semi-structured clinical interview including current medications and relevant comorbidities. Motor and non-motor symptoms were assessed using the MDS-UPDRS[1] which is comprised of four parts: patient and/or informant-rated non-motor experiences of daily living (Part I), motor experiences of daily living (Part II), clinician-rated motor examination (Part III), and motor complications (Part IV). Sleep disturbances were evaluated using the Scales for Outcomes in Parkinson's Disease-Sleep (SCOPA-Sleep).[2] The SCOPA-Sleep Nocturnal subscale provides an index of subjective nighttime sleep quality including sleep onset, maintenance and early waking. The SCOPA-Sleep Daytime subscale evaluates self-reported daytime somnolence and fatigue. Higher scores indicate greater disturbance on both subscales. Hallucinations were defined as a score ≥1 on MDS-UPDRS Part I item 2 ("Over the past week have you seen things that were not really there?"). The Psychosis and Hallucinations Questionnaire (PsycH-Q) provides a dimensional measure of hallucination severity and associated attentional and sleep disturbances across a broader psychotic phenotype, validated in both Parkinson's disease (PD)[3] and dementia with Lewy bodies (DLB).[4] Cognitive fluctuation severity in DLB was assessed via semi-structured interview with the patient and caregiver and rated on the Clinician Assessment of Fluctuations Scale (CAF),[5] which yields a composite score of fluctuation frequency and severity. The neuropsychological battery included the Montreal Cognitive Assessment (MoCA),[6] Trail Making Test Parts A and B,[7] and the Wechsler Memory Scale (Digit Span Forwards, Digit Span Backwards, and Logical Memory I and II).[8] Individual cognitive test scores were z-scored relative to the healthy control group mean and standard deviation prior to analysis. A reusable protocol is available at protocols.io (https://doi.org/10.17504/protocols.io.8epv5w336v1b/v1)

#### 1.2 MRI acquisition and preprocessing

Results included in this manuscript come from preprocessing performed using *fMRIPrep* 24.1.1[9,10]; (RRID:SCR_016216), which is based on *Nipype* 1.8.1[11,12]; (RRID:SCR_002502). A reusable protocol is available at protocols.io (https://doi.org/10.17504/protocols.io.8epv5w3b6v1b/v1).

##### 1.2.1 MRI acquisition

Both whole-brain structural T1-weighted and resting-state BOLD fMRI were obtained on a 3-Tesla scanner (General Electric) with the following acquisition parameters. Sagittal 3D T1-weighted structural images were acquired with TE = 2.7 ms, TR = 7.2 ms, flip angle = 12°, acquisition matrix 256 × 256, 200 slices, slice thickness = 1 mm. T2*-weighted echo-planar functional images were acquired with TE = 36 ms, TR = 3000 ms, flip angle = 90°, 32 contiguous axial slices covering the whole brain, raw voxel size = 3.75 × 3.75 × 3 $mm^3$.

##### 1.2.2 Anatomical data preprocessing

The T1-weighted (T1w) image was corrected for intensity non-uniformity (INU) using N4BiasFieldCorrection[13]; distributed with ANTs 2.3.1, (RRID:SCR_004757) and used as the T1w reference. The T1w reference was skull-stripped using the antsBrainExtraction.sh workflow (ANTs), with OASIS30ANTs as the target template. Brain tissue segmentation (CSF, WM, GM) was performed using fast[14]; FSL 6.0.3, (RRID:SCR_002823). Brain surfaces were reconstructed using "recon-all" with FreeSurfer 7.3.2[15], (RRID:SCR_001847), and the brain mask estimated previously was refined with a custom variation of the method to reconcile ANTs-derived and FreeSurfer-derived segmentations of the cortical gray-matter of Mindboggle[16] (RRID:SCR_002438). Volume-based spatial normalization to two standard spaces (MNI152NLin6Asym, MNI152NLin2009cAsym) was performed through nonlinear registration with antsRegistration (ANTs 2.5.3), using brain-extracted versions of both T1w reference and the T1w template. The following templates were selected for spatial normalization and accessed with TemplateFlow 24.2.0[17]: FSL's MNI ICBM 152 non-linear 6th Generation Asymmetric Average Brain Stereotaxic Registration Model[18] (RRID:SCR_002823; TemplateFlow ID: MNI152NLin6Asym), ICBM 152 Nonlinear Asymmetrical template version 2009c[19] (RRID:SCR_008796; TemplateFlow ID: MNI152NLin2009cAsym). Grayordinate "dscalar" files containing 91k samples were resampled onto fsLR using the Connectome Workbench.[20]

##### 1.2.3 Functional data preprocessing

For each subject's resting-state BOLD run, the following preprocessing was performed. First, a reference volume was generated, using a custom methodology of fMRIPrep, for use in head motion correction. Head-motion parameters with respect to the BOLD reference (transformation matrices, and six corresponding rotation and translation parameters) are estimated before any spatiotemporal filtering using mcflirt[21] (FSL). The BOLD reference was then co-registered to the T1w reference using bbregister (FreeSurfer) which implements boundary-based registration.[22] Co-registration was configured with six degrees of freedom. BOLD runs were normalized to MNI152NLin6Asym and MNI152NLin2009cAsym spaces via antsApplyTransforms (ANTs) and mri_vol2surf (FreeSurfer).

Several confounding time-series were calculated based on the preprocessed BOLD: framewise displacement (FD), DVARS and three region-wise global signals. FD was computed using two formulations following Power et al.[23] (absolute sum of relative motions) and Jenkinson et al.[21] (relative root mean square displacement between affines). FD and DVARS are calculated for each functional run, both using their implementations in Nipype (following the definitions by Power et al.[23]). The three global signals are extracted within the CSF, the WM, and the whole-brain masks. Additionally, a set of physiological regressors were extracted to allow for component-based noise correction[24] (CompCor). Principal components are estimated after high-pass filtering the preprocessed BOLD time-series (using a discrete cosine filter with 128s cut-off) for the two CompCor variants: temporal (tCompCor) and anatomical (aCompCor). tCompCor components are then calculated from the top 2% variable voxels within the brain mask. For aCompCor, three probabilistic masks (CSF, WM and combined CSF+WM) are generated in anatomical space. The implementation differs from that of Behzadi et al. in that instead of eroding the masks by 2 pixels on BOLD space, a mask of pixels that likely contain a volume fraction of GM is subtracted from the aCompCor masks. This mask is obtained by dilating a GM mask extracted from the FreeSurfer's aseg segmentation, and it ensures components are not extracted from voxels containing a minimal fraction of GM. Finally, these masks are resampled into BOLD space and binarized by thresholding at 0.99 (as in the original implementation). Components are also calculated separately within the WM and CSF masks. For each CompCor decomposition, the *k* components with the largest singular values are retained, such that the retained components' time series are sufficient to explain 50 percent of variance across the nuisance mask (CSF, WM, combined, or temporal). The remaining components are dropped from consideration. The head-motion estimates calculated in the correction step were also placed within the corresponding confounds file. The confound time series derived from head motion estimates and global signals were expanded with the inclusion of temporal derivatives and quadratic terms for each.[25] Frames that exceeded a threshold of 0.5 mm FD or 1.5 standardized DVARS were annotated as motion outliers. Additional nuisance timeseries are calculated by means of principal components analysis of the signal found within a thin band (crown) of voxels around the edge of the brain.[26] Gridded (volumetric) resamplings were performed using nitransforms, configured with cubic B-spline interpolation.

Many internal operations of fMRIPrep use Nilearn 0.10.4[27] (RRID:SCR_001362), mostly within the functional processing workflow. For further details, please see fMRIPrep's documentation.

#### 1.3 Global BOLD and CSF signal extraction and denoising

##### 1.3.1 Gray matter mask

The gray matter mask used for global BOLD signal extraction was derived from the Harvard-Oxford cortical atlas[28] (threshold 25%, 2 mm resolution), warped from MNI152NLin6Asym standard space to each participant's native BOLD space (Fig S1). Warping was performed in two steps using fMRIPrep-generated transforms: the MNI-to-T1w nonlinear composite warp, followed by the BOLD reference-to-T1w co-registration transform, both applied via SimpleITK. The warped atlas mask was intersected with each participant's functional field of view to exclude out-of-brain voxels.

##### 1.3.2 Cerebrospinal fluid mask

The CSF inflow signal was extracted from a manually delineated mask placed at the most inferior slice of the raw functional image, where fluid entering the acquisition volume has had minimal exposure to radiofrequency pulses (Fig S1). CSF voxels were identified based on elevated signal intensity relative to surrounding tissue on T2*-weighted images. The CSF mask was intersected with the functional field of view prior to signal extraction.

##### 1.3.3 Denoising

Signal extraction and denoising were performed using Nilearn 0.10.4.[27] Voxelwise BOLD time series were extracted from the gray matter and CSF masks separately. Both signal timeseries underwent linear and quadratic detrending to remove slow signal drift, without additional motion parameter regression, consistent with Fultz et al.[29] and previous BOLD-CSF coupling studies.[30,31] Signals were then bandpass filtered (0.01-0.10 Hz) to isolate low-frequency fluctuations and exclude high-frequency physiological noise. Global BOLD and CSF signals were computed as the spatial mean across all voxels within their respective mask. BOLD and CSF timeseries were z-scored prior to coupling analyses. As illustrated in Figure S1, changes in the global BOLD signal are associated with changes in the CSF inflow signal, reflecting the coupling of cerebrovascular dynamics with pulsatile CSF displacement.

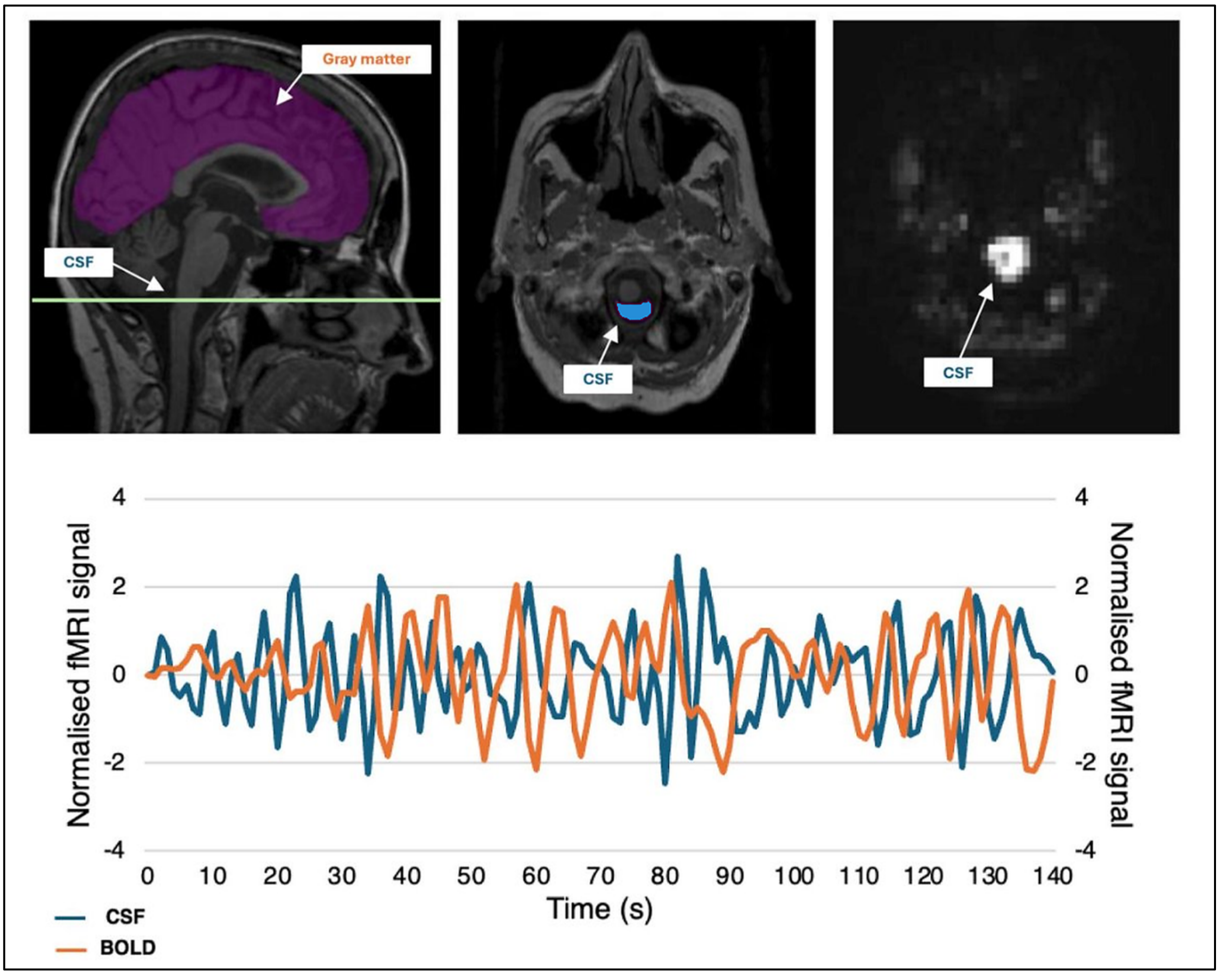

**Figure S1. Gray matter and CSF mask placement and representative signal timeseries.**

The gray matter mask (purple) was derived from the Harvard-Oxford cortical and subcortical atlas (25% threshold) warped to native BOLD space, with the green line indicating the axial slice used for CSF mask placement (left panel). The CSF mask (blue) was placed at the most inferior slice of the raw functional image, where inflow-related signal intensity changes are maximal (middle panel). The corresponding bright signal intensity of the CSF at this slice is visible on the raw T2*-weighted functional image (right panel). The lower panel shows the normalised global BOLD (orange) and CSF inflow (blue) timeseries from a representative control subject.

#### 1.4 Basal forebrain and brainstem segmentation

Basal forebrain volumes were extracted using ScLimbic[32] (mri_sclimbic_seg), a deep learning segmentation tool integrated into FreeSurfer. The tool uses a U-Net architecture trained on 39 manually labelled T1-weighted datasets with spatial, intensity, contrast and noise augmentation to achieve contrast-agnostic segmentation of several subcortical limbic structures including the basal forebrain, nucleus accumbens, septal nuclei, hypothalamus, mammillary bodies and fornix. Basal forebrain volume in $mm^3$ was extracted from segmentation outputs. Quality control was performed using the built-in z-score and posterior confidence metrics.

Brainstem ascending arousal network (AAN) nuclei were segmented using AANSegment,[33] an automated segmentation tool released for integrated use with FreeSurfer. This tool uses a probabilistic atlas of 10 AAN nuclei constructed from manual tracings of *ex vivo* diffusion MRI scans acquired in five human brain specimens at 750 µm isotropic resolution, with neuroanatomic boundaries annotated using 200 µm 7T MRI and nucleus-specific immunostaining. The Bayesian segmentation algorithm deforms this atlas to identify each nuclei in a resolution- and contrast-adaptive manner, enabling application to standard *in vivo* T1-weighted MRI. Whole brainstem AAN composite volume in $mm^3$ was extracted from segmentation outputs.

Total gray matter volume was obtained from FreeSurfer summary statistics (aseg.stats). Bilateral striatal volume was derived from FreeSurfer aseg labels by summing the left and right caudate, putamen and nucleus accumbens volumes.

To account for inter-individual variation in head size, basal forebrain, brainstem, striatal and total gray matter volumes were adjusted for estimated total intracranial volume (eTIV), derived from the FreeSurfer output using the residual correction method.[34] A regression of raw region of interest volume on eTIV was fitted across all participants, and the adjusted volume was computed as the residual plus the grand mean. All segmentations were performed in native T1-weighted space and visually inspected using FreeView.

### 2. Supplementary results

#### 2.1 Clinical and cognitive regression analyses

**Table S1. Association between global BOLD-CSF coupling strength and clinical symptom severity**

| Measure | Standardized β | 95% CI | *p* | $p_{FDR}$ |
|---|---|---|---|---|
| Hallucinations | 0.533 | [0.306, 0.754] | 0.005 | 0.015 |
| SCOPA-Sleep Night | 0.631 | [0.257, 0.843] | 0.001 | 0.005 |
| SCOPA-Sleep Day | 0.109 | [-0.353, 0.560] | 0.593 | 0.712 |
| MDS-UPDRS I | 0.183 | [-0.239, 0.564] | 0.360 | 0.540 |
| MDS-UPDRS II | 0.326 | [-0.074, 0.638] | 0.098 | 0.196 |
| MDS-UPDRS III | 0.003 | [-0.331, 0.379] | 0.989 | 0.989 |

Standardized β and p values from permutation-based regression (two-sided, 10000 permutations). MDS-UPDRS, Movement Disorder Society Unified Parkinson's Disease Rating Scale; SCOPA-Sleep, Scale for Outcomes in Parkinson's Disease-Sleep. Hallucinations defined by a score ≥1 on MDS-UPDRS Part I item 2.

**Table S2. Jackknife leave-one-out sensitivity analyses for exploratory subset analyses**

| Measure | Observed β (std) | LOO β (std) range | Direction consistency | *p* |
|---|---|---|---|---|
| CAF | 0.680 | [0.529, 0.769] | 100% | 0.037 |
| PsycH-Q | 0.590 | [0.510, 0.656] | 100% | 0.025 |

LOO, leave-one-out; observed β (std), standardised regression coefficient from the full model; LOO β (std) range, minimum and maximum standardised β across all LOO iterations (n iterations = sample size); direction consistency, proportion of LOO iterations in which the sign of β agreed with the full model; *p*, two-tailed permutation *p*-value from the full model (10000 permutations). CAF, Clinical Assessment of Fluctuations; PsycH-Q, Psychosis and Hallucinations Questionnaire

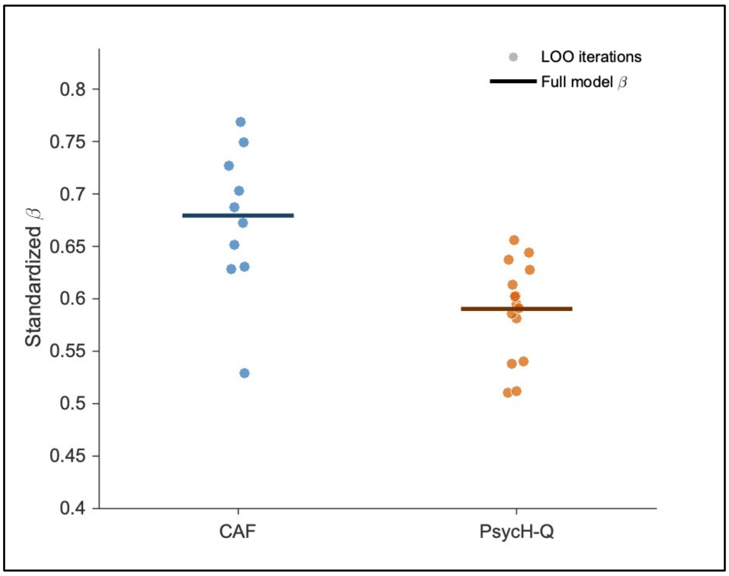

**Figure S2. Plot of Jackknife leave-one-out sensitivity analysis**

Each point represents the standardized regression coefficient (β) obtained from a model with one participant removed. The horizontal line indicates the full model estimate. All leave-one-out iterations were positive and consistent in direction for both the Clinician Assessment of Fluctuations (CAF; n = 10) and Psychosis and Hallucinations Questionnaire (PsycH-Q; n = 15), indicating that associations were not driven by single influential observations.

**Table S3. Association between global BOLD-CSF coupling strength and cognitive performance**

**Whole sample**

| **Measure** | **Standardized β** | **95% CI** | ***p*** | $p_{FDR}$ |
|---|---|---|---|---|
| MoCA | -0.153 | [-0.473, 0.370] | 0.723 | 0.855 |
| Digit Span Forwards | -0.332 | [-0.610, -0.012] | 0.059 | 0.138 |
| Digit Span Backwards | -0.577 | [-0.754, -0.367] | 0.001 | 0.004 |
| Trails A | 0.036 | [-0.380, 0.415] | 0.840 | 0.855 |
| Trails B | -0.033 | [-0.387, 0.325] | 0.855 | 0.855 |
| Logical Memory I | -0.416 | [-0.705, -0.035] | 0.015 | 0.053 |
| Logical Memory II | -0.226 | [-0.587, 0.255] | 0.196 | 0.343 |

**LBD only**

| **Measure** | **Standardized β** | **95% CI** | ***p*** | $p_{FDR}$ |
|---|---|---|---|---|
| MoCA | -0.039 | [-0.429, 0.497] | 0.852 | 0.852 |
| Digit Span Forwards | -0.296 | [-0.649, 0.099] | 0.143 | 0.334 |
| Digit Span Backwards | -0.550 | [-0.782, -0.251] | 0.005 | 0.035 |
| Trails A | 0.167 | [-0.274, 0.542] | 0.430 | 0.602 |
| Trails B | 0.162 | [-0.204, 0.514] | 0.428 | 0.602 |
| Logical Memory I | -0.374 | [-0.730, 0.090] | 0.063 | 0.221 |
| Logical Memory II | -0.096 | [-0.514, 0.433] | 0.640 | 0.747 |

Standardized β and p values from permutation-based regression (two-sided, 10000 permutations). LBD, Lewy body disorder (combined PD and DLB).

### 2.2 Regional BOLD-CSF coupling

**Table S4. Group differences in network-level BOLD-CSF coupling strength between controls and combined LBD group**

| Network | Control adj. mean | LBD adj. mean | Hedges' $g$ | $p$ | $p_{FDR}$ |
|---|---|---|---|---|---|
| Visual | -0.268 | -0.134 | 0.755 | 0.016 | 0.044 |
| Somatomotor | -0.215 | -0.081 | 0.737 | 0.019 | 0.044 |
| Dorsal attention | -0.223 | -0.094 | 0.719 | 0.018 | 0.044 |
| Ventral attention | -0.132 | -0.081 | 0.433 | 0.158 | 0.184 |
| Frontoparietal | -0.181 | -0.082 | 0.627 | 0.042 | 0.074 |
| Default mode | -0.160 | -0.101 | 0.454 | 0.142 | 0.184 |
| Limbic | -0.096 | -0.086 | 0.089 | 0.778 | 0.778 |

Adjusted (adj.) means, Hedges' $g$ and $p$ values derived from covariate-adjusted (age, sex) non-parametric permutation testing (two-sided, 10000 permutations) comparing the combined LBD group (PD and DLB) to controls. DLB, dementia with Lewy bodies; LBD, Lewy body disorder; PD, Parkinson's disease

#### 2.3 Volumetric regression analyses

**Table S5. Association between basal forebrain and brainstem volumes and BOLD-CSF coupling strength**

**Whole sample - adjusted for age and sex**

| Predictor | Outcome | Standardized β | 95% CI | *p* | $p_{FDR}$ |
|---|---|---|---|---|---|
| Basal Forebrain | Global | -0.085 | [-0.378, 0.223] | 0.617 | 0.617 |
| | Unimodal | -0.089 | [-0.421, 0.262] | 0.591 | 0.617 |
| | Attentional | -0.171 | [-0.467, 0.149] | 0.301 | 0.452 |
| Brainstem | Global | -0.316 | [-0.546, -0.037] | 0.052 | 0.104 |
| | Unimodal | -0.348 | [-0.559, -0.093] | 0.031 | 0.093 |
| | Attentional | -0.358 | [-0.561, - 0.124] | 0.027 | 0.093 |

**Whole sample - unadjusted**

| Predictor | Outcome | Standardized β | 95% CI | *p* | $p_{FDR}$ |
|---|---|---|---|---|---|
| Basal Forebrain | Global | -0.384 | [-0.626, -0.101] | 0.016 | 0.019 |
| | Unimodal | -0.375 | [-0.623, -0.098] | 0.019 | 0.019 |
| | Attentional | -0.459 | [-0.676, -0.199] | 0.003 | 0.008 |
| Brainstem | Global | -0.431 | [-0.636, -0.182] | 0.006 | 0.009 |
| | Unimodal | -0.456 | [-0.647, -0.238] | 0.004 | 0.008 |
| | Attentional | -0.494 | [-0.677, -0.282] | 0.001 | 0.006 |

**LBD only - adjusted for age and sex**

| Predictor | Outcome | Standardized β | 95% CI | *p* | $p_{FDR}$ |
|---|---|---|---|---|---|
| Basal Forebrain | Global | -0.156 | [-0.498, 0.201] | 0.442 | 0.456 |
| | Unimodal | -0.148 | [-0.543, 0.283] | 0.456 | 0.456 |
| | Attentional | -0.239 | [-0.581, 0.135] | 0.224 | 0.336 |
| Brainstem | Global | -0.304 | [-0.566, 0.041] | 0.130 | 0.260 |
| | Unimodal | -0.319 | [-0.554, 0.003] | 0.108 | 0.260 |
| | Attentional | -0.354 | [-0.613, -0.050] | 0.070 | 0.260 |

**LBD only - unadjusted**

| Predictor | Outcome | Standardized β | 95% CI | *p* | $p_{FDR}$ |
|---|---|---|---|---|---|
| Basal Forebrain | Global | -0.458 | [-0.725, -0.106] | 0.018 | 0.018 |
| | Unimodal | -0.462 | [-0.744, -0.120] | 0.018 | 0.018 |
| | Attentional | -0.521 | [-0.748, -0.197] | 0.005 | 0.015 |
| Brainstem | Global | -0.451 | [-0.673, -0.146] | 0.018 | 0.018 |
| | Unimodal | -0.485 | [-0.697, -0.224] | 0.010 | 0.018 |
| | Attentional | -0.523 | [-0.716, -0.272] | 0.004 | 0.015 |

Standardized β and *p* values from permutation-based regression (two-sided, 10000 permutations), false discovery rate (FDR) corrected across the six comparisons within each sample and adjustment level. LBD, Lewy body disorder (combined PD and DLB).

**Table S6. Association between total gray matter volume and striatal volume and BOLD-CSF coupling strength**

**Whole sample - adjusted for age and sex**

| **Predictor** | **Outcome** | **Standardized β** | **95% CI** | ***p*** | $p_{FDR}$ |
|---|---|---|---|---|---|
| Total gray matter volume | Global | -0.402 | [-0.711, 0.016] | 0.050 | 0.449 |
| | Unimodal | -0.297 | [-0.623, 0.110] | 0.164 | 0.493 |
| | Attentional | -0.312 | [-0.629, 0.078] | 0.131 | 0.493 |
| Striatum | Global | -0.048 | [-0.318, 0.240] | 0.742 | 0.969 |
| | Unimodal | -0.107 | [-0.376, 0.186] | 0.466 | 0.969 |
| | Attentional | -0.005 | [-0.299, 0.265] | 0.969 | 0.969 |

**Whole sample - unadjusted**

| **Predictor** | **Outcome** | **Standardized β** | **95% CI** | ***p*** | $p_{FDR}$ |
|---|---|---|---|---|---|
| Total gray matter volume | Global | -0.517 | [-0.727, -0.251] | 0.001 | 0.005 |
| | Unimodal | -0.457 | [-0.692, -0.164] | 0.004 | 0.011 |
| | Attentional | -0.513 | [-0.740, -0.235] | 0.001 | 0.005 |
| Striatum | Global | -0.081 | [-0.362, 0.189] | 0.625 | 0.703 |
| | Unimodal | -0.134 | [-0.413, 0.159] | 0.406 | 0.522 |
| | Attentional | -0.031 | [-0.330, 0.250] | 0.853 | 0.853 |

**LBD only - adjusted for age and sex**

| **Predictor** | **Outcome** | **Standardized β** | **95% CI** | ***p*** | $p_{FDR}$ |
|---|---|---|---|---|---|
| Total gray matter volume | Global | -0.530 | [-0.898, -0.025] | 0.049 | 0.220 |
| | Unimodal | -0.375 | [-0.733, 0.049] | 0.172 | 0.516 |
| | Attentional | -0.523 | [-0.852, 0.038] | 0.047 | 0.220 |
| Striatum | Global | 0.019 | [-0.295, 0.429] | 0.922 | 0.947 |
| | Unimodal | -0.012 | [-0.356, 0.433] | 0.947 | 0.947 |
| | Attentional | 0.152 | [-0.185, 0.508] | 0.393 | 0.885 |

**LBD only - unadjusted**

| **Predictor** | **Outcome** | **Standardized β** | **95% CI** | ***p*** | $p_{FDR}$ |
|---|---|---|---|---|---|
| Total gray matter volume | Global | -0.583 | [-0.801, -0.278] | 0.001 | 0.005 |
| | Unimodal | -0.555 | [-0.785, -0.193] | 0.002 | 0.005 |
| | Attentional | -0.631 | [-0.838, -0.323] | <0.001 | 0.004 |
| Striatum | Global | -0.010 | [-0.338, 0.347] | 0.960 | 0.960 |
| | Unimodal | -0.052 | [-0.403, 0.305] | 0.797 | 0.897 |
| | Attentional | 0.106 | [-0.219, 0.402] | 0.604 | 0.776 |

Standardized β and *p* values from permutation-based regression (two-sided, 10000 permutations), false discovery rate (FDR) corrected across the six comparisons within each sample and adjustment level. LBD, Lewy body disorder (combined PD and DLB).

### 2.4 Control analyses

**Table S7. Group differences in motion across diagnostic groups**

| Measure | Contrast | Hedges' *g* | *p* |
|---|---|---|---|
| Mean FD | Control vs PD | 0.300 | 0.439 |
| | Control vs DLB | 0.200 | 0.640 |
| | PD vs DLB | -0.142 | 0.731 |
| Proportion high-motion volumes | Control vs PD | 0.277 | 0.482 |
| | Control vs DLB | -0.046 | 0.918 |
| | PD vs DLB | -0.338 | 0.427 |

High motion volumes were defined as framewise displacement (FD) >0.5mm or DVARS >1.5. Hedges' *g* and *p* values derived from covariate-adjusted (age, sex) non-parametric permutation testing (two-sided, 10000 permutations). DLB, dementia with Lewy bodies; FD, framewise displacement; PD, Parkinson's disease

**Table S8. Group differences in BOLD-CSF coupling controlling for age, sex and mean framewise displacement**

| BOLD-CSF coupling measure | Contrast | Hedges' *g* | *p* | $p_{FDR}$ |
|---|---|---|---|---|
| Global | Control vs PD | 0.790 | 0.022 | 0.033 |
| | Control vs DLB | 0.695 | 0.021 | 0.033 |
| | PD vs DLB | -0.284 | 0.383 | 0.383 |
| Unimodal | Control vs LBD | 0.841 | 0.007 | 0.021 |
| | PD vs DLB | -0.269 | 0.405 | 0.609 |
| Attentional | Control vs LBD | 0.623 | 0.044 | 0.066 |
| | PD vs DLB | -0.098 | 0.765 | 0.765 |
| Transmodal | Control vs LBD | 0.493 | 0.102 | 0.102 |
| | PD vs DLB | -0.272 | 0.406 | 0.609 |

Hedges' *g* and *p* values derived from covariate-adjusted (age, sex, head motion) non-parametric permutation testing (two-sided, 10000 permutations). DLB, dementia with Lewy bodies; LBD, Lewy body disorder (combined patient group); PD, Parkinson's disease

**Table S9. Associations between levodopa equivalent daily dose and BOLD-CSF coupling strength**

| Sample | Outcome | Standardized β | *p* |
|---|---|---|---|
| PD only (n = 17) | Global | 0.109 | 0.679 |
| | Unimodal | 0.036 | 0.892 |
| | Attentional | 0.129 | 0.626 |
| | Transmodal | 0.223 | 0.387 |
| Combined PD and DLB on levodopa (n = 21) | Global | 0.110 | 0.640 |
| | Unimodal | 0.051 | 0.828 |
| | Attentional | 0.124 | 0.585 |
| | Transmodal | 0.195 | 0.401 |

PD only model adjusted for age and sex; combined model additionally adjusted for diagnosis. Standardized β and *p* values from permutation-based regression (two-sided, 10000 permutations) DLB, dementia with Lewy bodies; PD, Parkinson's disease

| RESOURCE TYPE | RESOURCE NAME | SOURCE | IDENTIFIER | NEW/REUSE | ADDITIONAL INFORMATION |
|---|---|---|---|---|---|
| **Dataset** | De-identified derived data | GitHub/Zenodo edit link | https://doi.org/10.5281/zenodo.21897287 | New | Clinical and cognitive data; whole brain and network level BOLD-CSF coupling values; total intracranial volume, total gray matter volume and regional volumes; includes data dictionary. https://github.com/annaignatavicius/BOLD_CSF_LBD/tree/main/data |
| **Code** | Analysis scripts | GitHub/Zenodo edit link | https://doi.org/10.5281/zenodo.21897287 | New | Code for signal extraction and denoising; quantification of global and parcel-wise regional BOLD-CSF coupling); includes README. https://github.com/annaignatavicius/BOLD_CSF_LBD/tree/main/code |
| **Software** | fMRIPrep v24.1.1 | Poldrack Lab Stanford | https://doi.org/10.1038/s41592-018-0235-4 (RRID:SCR_016216) | Reuse | |
| **Software** | Python v3.10 | Python Software Foundation | https://www.python.org/ (RRID:SCR_008394) | Reuse | |
| **Software** | FreeSurfer v7.3.2 | Athinoula A. Martinos Center for Biomedical Imaging | http://surfer.nmr.mgh.harvard.edu/ (RRID:SCR_001847) | Reuse | |
| **Software** | MATLAB R2023a | MathWorks | https://www.mathworks.com/ (RRID:SCR_001622) | Reuse | |
| **Protocol** | Clinical and neuropsychological assessment in Lewy body disorders | protocols.io | https://doi.org/10.17504/protocols.io.8epv5w336v1b/v1 | New | |
| **Protocol** | Structural and resting-state functional MRI acquisition and preprocessing | protocols.io | https://doi.org/10.17504/protocols.io.8epv5w3b6v1b/v1 | New | |
